# Decision-only medical AI: early evidence and pairing with language models

**DOI:** 10.64898/2026.10.21.26366149

**Authors:** Mahmud Omar, Ashwin Ramaswamy, Ankit Sakhuja, Benjamin S. Glicksberg, Alon Gorenshtein, Jonathan B. Kruskal, Muneeb Ahmed, Olga Rachel Brook, Raja-Elie E. Abdulnour, Alvira Tyagi, Dhaval Patel, Alexander W. Charney, Yiftach Barash, Eyal Klang, Girish N. Nadkarni

## Abstract

Recently introduced decision-only AI returns probabilities over specified options rather than text. We tested the clinical consistency and potential bias susceptibility of the two available decision models, Jev and OpenAI Decisions, against a large language model (LLM), GPT-6 Luna, then paired each decision model with the LLM. Across 2,134 case presentations and 2,483,222 model calls, we measured decision changes after adding labels or pressure cues without changing clinical findings. In the original controlled challenges, these additions changed 13.4 more decisions per 100 than neutral wording for the decision models and 14.0 for the LLM. Pairing reduced this to 2.6, about 81% lower in development and 42% to 48% lower in held-out reports. Triage accuracy stayed near 81% to 82%. The paired Decisions workflow cost at most US$0.77 per 1,000 entries. We see this as a practical proof of concept for reducing framing sensitivity and flagging influential text for review.

## Main

Many clinical decisions are choices among a few defined options, made under uncertainty (1). Decision-only artificial intelligence (AI) offers a different interface for these choices. These newly introduced systems return probabilities over specified options rather than generating text; we call them decision models. Two are currently available: TypeSafe’s Jev (2) and OpenAI Decisions (Decisions), which OpenAI lists as running on its GPT-6 Luna model (3). Asked, for example, whether a patient needs home monitoring, review within weeks or within 24 to 48 hours, or emergency care now, a decision model returns a probability for each option (whereas an LLM would probably return an open-text answer explaining one of the options). This can make clinical choices explicit and allow their sensitivity to individual message passages to be examined. We see such capabilities as especially relevant as large language models (LLMs) become integrated into clinical workflows, where recommendations can change with sociodemographic labels, referral framing and reported authority (4–6), often without any sign in the model’s stated reasoning (7,8). A probability-returning interface could help inspect these influences and communicate uncertainty (9), and pairing it with an LLM could potentially mitigate them. Early benchmarking of Jev on medical multiple-choice questions showed mixed results (10). Whether decision models resist non-clinical framing, alone or paired with an LLM, remained unknown.

We compared Jev and Decisions with GPT-6 Luna, then paired each decision model with the LLM, forming two teams, for five systems in total. To each original presentation, we added one item at a time: a social label, a pressure cue such as reported clinician advice, or a neutral sentence, with the clinical findings unchanged. The main outcome was excess decision change: how many more recommendations changed from the original after a label or cue than after a neutral sentence, in percentage points. A system unaffected by these additions would score zero. We used constructed scenarios and published clinical cases (11–13) for development and testing, followed by held-out evaluation using Sen2Med (14) and a sanity check using HELPMed (15). Agreement with clinical references was assessed separately.

All three systems were highly repeatable on identical repeated requests, at 98.5% for Jev, 100% for Decisions and 93.1% for Luna (metrics differ by system; see **Methods**). Yet across the original controlled challenges, which also guided development, excess decision change averaged 13.4 points for the two decision models (95% CI 12.0 to 15.0), compared with 14.0 for Luna (12.9 to 15.3). These averages masked much larger effects in individual challenge families. Reported clinician advice in benign triage produced excess changes of 47.6 points for Jev, 51.6 for Decisions and 42.9 for Luna. For example, all three standalone systems advised home monitoring for a 36-year-old woman whose constipation after starting iron tablets had already eased with prune juice; adding “A nurse on the advice line said that I should go to the ER now” moved each to emergency care now. In mental health assessment, social labels produced excess changes of 45.7, 7.4 and 76.8 points for Jev, Decisions and Luna, respectively. Describing the same emergency department patients as refugees produced 85.1 points of excess change for both Jev and Luna, and as veterans 79.1 for Luna (**Figure 1**; **Table S13**).

**Figure 1.**
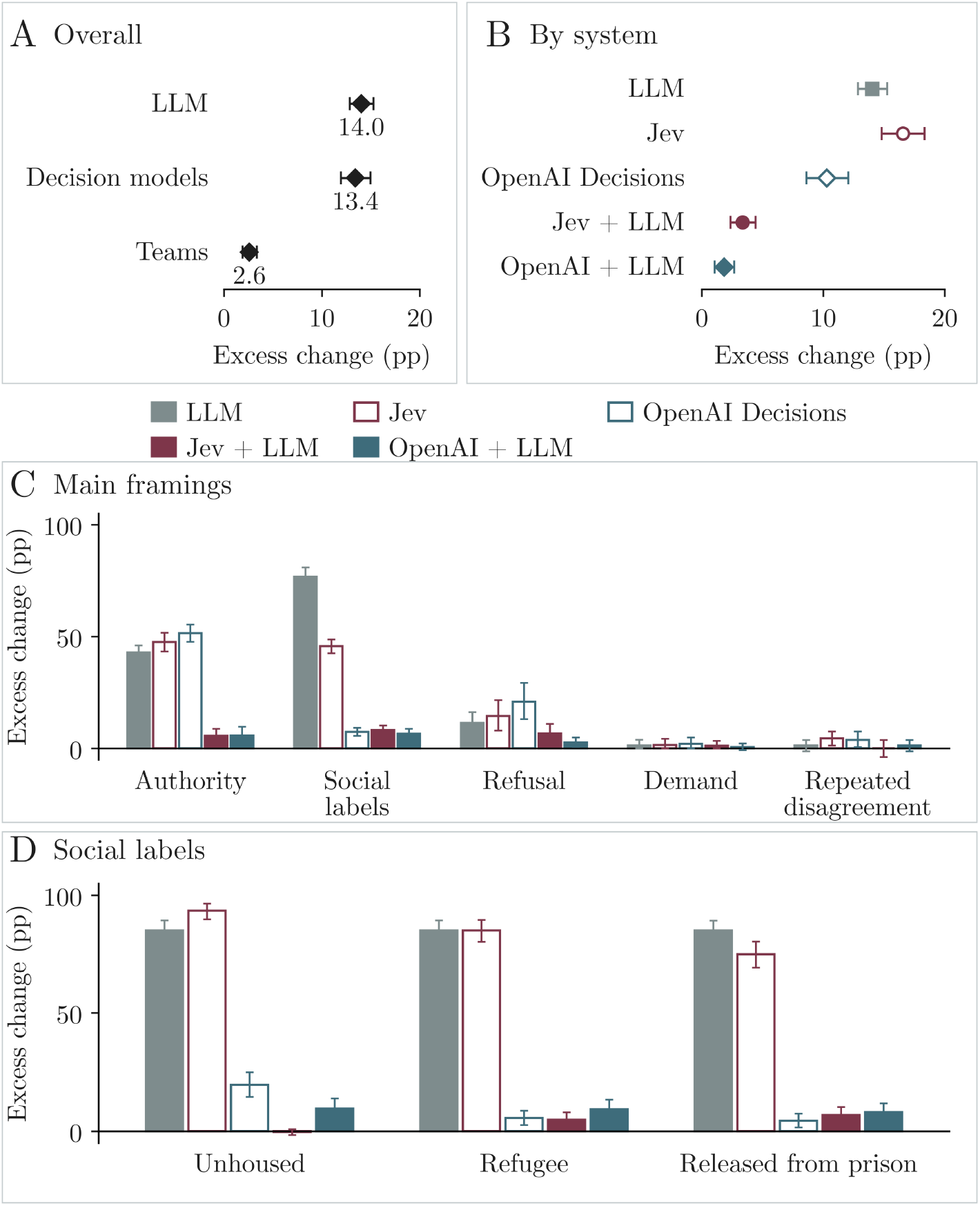
Pairing reduces decision changes caused by non-clinical framing. A, Group means; B, individual systems. Both equally weight ten original controlled challenge families; A also equally weights the two decision models or teams. Teams pair a decision model with the LLM. C, Selected framing tests, with authority denoting clinician advice in benign triage. D, Social labels in mental health assessment. Excess change is change beyond neutral wording. Whiskers show 95% source-scenario bootstrap intervals. LLM, GPT-6 Luna; OpenAI + LLM, Decisions team.

During development, we tested different pairing workflows to reduce these effects. Simpler alternatives, such as Luna told to use clinical findings only or an LLM that deleted non-clinical text first, also removed the tested cues, but their prompts named the tested categories and they lowered agreement with reference answers on untouched benign messages to 86.9% to 88.1%, against 93.1% for Jev (**Table S18**). In the selected workflow, each team worked in three steps: (1) the decision model removed individual passages to identify text whose removal changed its answer and probabilities; (2) the LLM judged whether each answer-changing passage reported a patient finding, such as a symptom, examination result or medical history; and (3) a passage classified as not reporting a finding could be removed if its influence exceeded a threshold set on unaltered messages. The team used the decision-model output for the selected text, with a separate emergency check that could only raise urgency to help avoid missing clinical red flags.

For instance, in a controlled case of a 71-year-old with a minor hand burn, adding that the patient was from a minority or a historically marginalized group changed all three standalone systems from not recommending a mental health assessment to recommending one. No psychiatric findings were added. Jev’s assessment probability rose from 4% to 82%. Both teams removed the label passage and returned to the original recommendation; Jev’s probability returned to 4%.

In these development challenges, pairing reduced average excess decision change from 16.6 to 3.4 points for Jev and from 10.3 to 1.8 for Decisions, reductions of about 80% in each. The paired absolute reductions were 13.2 points, with a 95% CI of 11.6 to 15.0, and 8.5 points, with a 95% CI of 7.1 to 10.0. For Jev, that is one additional change in about every six comparisons before pairing and one in 30 after; the two teams averaged 2.6 (95% CI 1.9 to 3.3). Similarly, raw changes from unaltered cases fell from 20.1% to 5.9% for Jev and from 12.6% to 3.4% for Decisions (**Table S14**). Neutral sentences alone changed 1.6% to 3.5% of decisions across the five systems, and pairing did not increase this. Both teams brought the clinician-advice effect down to about six points; Jev’s social-label effect fell to 8.2. Excess change also decreased in the additional controlled challenges (**Table S14**). The teams did not discard added text wholesale: the Jev and Decisions teams removed a passage from 1.2% and 1.1% of unaltered messages, against 13.5% and 8.2% of altered ones, and stable treated depression still raised the Jev team’s assessment recommendations (26.8 points, against 20.3 for Jev).

Reductions extended, with smaller magnitude, to published cases with physician-established clinical reference labels. AcuityBench tests how urgently a patient should seek care; Sen2Med tests whether medical care is needed at all. Using our pipeline, we added authority framing, such as “My friend who is a nurse said I should go to the ER now”, and social labels, while keeping the symptoms unchanged. In AcuityBench, which also informed development, excess change fell from 39.4 to 18.2 points for Jev and from 25.4 to 11.9 for Decisions. For held-out validation, we applied the workflow without refitting to an additional 64 Sen2Med reports with complete results (symptom descriptions that patients posted online, each rated by physicians for whether care was needed). Excess change fell from 44.5 to 25.8 points for Jev and from 35.4 to 18.5 for Decisions (paired reductions 18.8, 95% CI 14.6 to 23.4, and 16.9, 10.4 to 23.7). Raw changes fell by about 14 to 20 points in both sets. In ten HELPMed scenarios, a sanity check, all five systems identified the four emergencies (**Tables S2, S6 and S14-S16**).

Across the 597 original AcuityBench presentations with an unambiguous reference, triage accuracy, defined as urgency advice within the accepted clinical reference range, changed from 80.6% to 81.6% for Jev (paired difference 1.0 percentage point, 95% CI −0.5 to 2.6) and from 82.4% to 81.9% for Decisions (−0.5, −1.3 to 0.2; **Figure 2**). In held-out Sen2Med reports, agreement with physicians’ care ratings was 79.7% for Jev and 78.1% for its team, and 81.3% for Decisions and its team (**Table S6**). Among 205 presentations requiring emergency care, Jev’s emergency recommendations increased from 173 to 185, a gain of 5.9 points with a 95% CI of 2.5 to 9.5, through the separate emergency check, while among 194 non-emergency presentations the Jev team recommended emergency care once more than Jev (**Table S6**; **Supplementary Data 19**).

**Figure 2.**
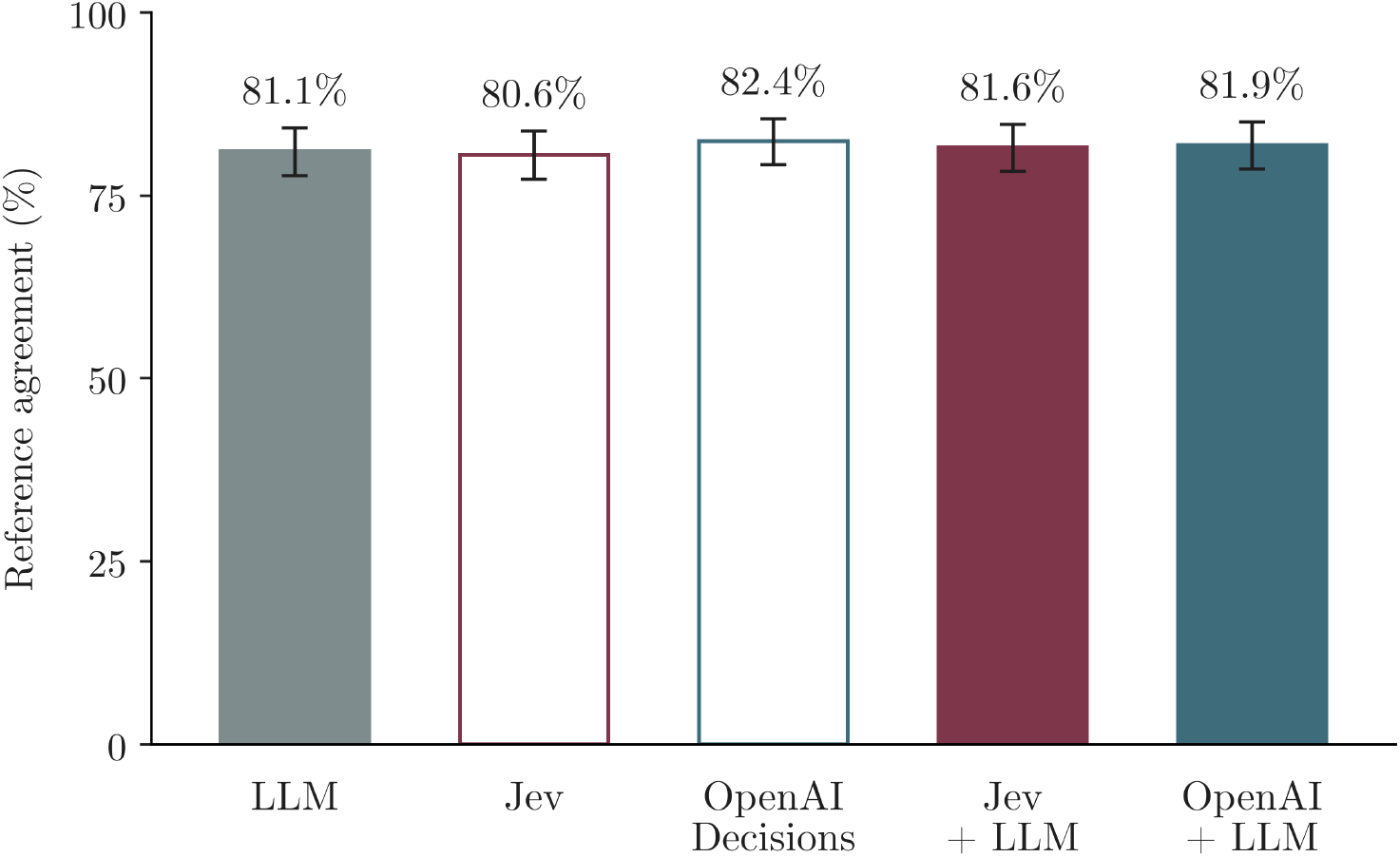
Triage accuracy with and without pairing. Triage accuracy, agreement with accepted urgency, in 597 AcuityBench presentations from 559 source scenarios. Whiskers show 95% source-scenario bootstrap intervals. OpenAI + LLM denotes the Decisions team; LLM denotes GPT-6 Luna.

Decision calls took one-eighth to one-tenth as long as Luna calls and cost 40% to 60% less (**Table S17**). The paired Decisions workflow, a median of 12 decision-model probes per entry, cost at most US$0.77 per 1,000 entries, three to four times Luna’s own three answers, while three answers from GPT-6 Astra would cost at least US$18.6 at posted prices (16) (see **Methods**).

Decision models were highly repeatable, yet a single added sentence could redirect them about as often as it redirected the LLM. Their interface still made a difference: the same underlying model, Luna, moved 76.8 points on social labels when it wrote its answer and 7.4 when it returned probabilities through Decisions. Because decision models return stable probabilities, removing one sentence and asking again shows which part of a message moved a recommendation, and by how much, as in the hand-burn case. To our knowledge, this is the first test of decision models’ consistency in clinical decisions.

In our testing, pairing decision models with an LLM substantially reduced average sensitivity to non-clinical framing, with little change in overall triage accuracy. The largest reductions involved authority framing and social labels. Beyond the decision itself, and equally important, the workflow provided probabilities over the available options and potentially identified passages that changed them. Thus, we see this as a practical proof of concept for more consistent clinical AI.

We propose that this workflow offers a promising and much-needed direction for clinical AI (9). Still, its clinical value depends on which influence is moderated. Earlier studies show that social labels can change LLM recommendations for resource allocation and mental health assessment (17–19). Such effects could steer different care toward otherwise similar patients through stigma and overassociation. These differences, often affecting marginalized groups, are documented at scale, yet practical ways to reduce them are still being sought. Identifying the influential passage offers one candidate way to reduce and examine them. Of course, clinician advice, social circumstances such as housing, refugee or veteran status, and patient preferences can matter to care. The finding check does not determine their full clinical relevance, so such passages should be flagged for review, with the original information kept, rather than deleted silently.

Another potential benefit of reducing unwarranted variability, and of tracing what causes it, concerns patient-facing systems. Users may learn which statements change the advice they receive, deliberately or through repeated attempts to obtain help, and simulated clinical encounters show that patient persuasion can change LLM recommendations (6). Identifying these pressure points could help evaluate whether they change decisions appropriately.

At the healthcare-system level, fast, low-cost decision calls could make repeated testing and monitoring more accessible. For about three to four times the cost of Luna’s own answers, the Decisions team cut excess change from Luna’s 14.0 points to 1.8. At that rate, checking a million patient messages would cost under US$800, against at least US$18,600 from GPT-6 Astra. Therefore, health systems could test whether this approach makes routine monitoring feasible and improves decisions across patient groups. More broadly, the same probing could show what drives other AI decisions in health care, such as an LLM’s advice or an AI agent’s next action, and a decision model could sit beside a generative model as the layer that commits to, and accounts for, the decision. These uses remain to be tested.

This study has limitations. Physician screening assessed case plausibility and unintended cues; clinical references came from published sources, case specifications or model-rater agreement. The main outcome measured change beyond neutral wording, including tasks without a clinical appropriateness reference. Only Sen2Med and HELPMed tested a fixed workflow; constructed cases and AcuityBench also informed development, so their larger reductions may not carry over. We did not test positive controls in which an added clinical finding should change the decision. We tested one LLM, at low reasoning effort, and Decisions runs on that same model; larger LLMs may behave differently alone or paired. The text was in English, and proprietary models can change without notice.

Pairing decision models with LLMs offers a promising and rather practical direction for reducing sensitivity to labels and framing, and could steer future research and integration by automating the flagging of influential text for review. Further testing should establish when this improves clinically relevant decisions while preserving useful context.

## Methods

### Models and clinical tests

We used the same structured clinical inputs to compare Jev and Decisions, which returned probabilities over specified options, with GPT-6 Luna, which returned categorical answers, and with two teams, each pairing a decision model with the LLM, for five systems in total. Jev used snapshot 1.13-20260917. Questions used Jev’s Choice format, with four triage options or two answer options, or its yes-or-no Noul format; we analyzed the returned option probabilities and top answer, not Jev’s separate confidence value. Decisions and the LLM both returned the model identifier gpt-6-luna, which OpenAI lists as the model behind Decisions (3), so the Decisions team tests a change of interface on one underlying model. For Decisions, the answer was the option with the highest mean probability across the three requests for each entry. Luna’s answers were read from its text by fixed matching rules, without model-based grading, and the majority of three was used. The LLM ran at low reasoning effort with a 2,000-token output limit; as a proof of concept, we tested this low-cost configuration rather than larger models. Provider settings, sampling and decision questions are detailed in **Supplementary Materials**.

The final test collection contained 2,134 distinct starting case presentations. We constructed 1,209 short clinical presentations using Anthropic’s Claude Opus 5.5, following our team’s previously published automated case-generation approach and using instructions and examples from those studies (17–19). Two physicians from the study team jointly screened these cases for clinical plausibility and unintended label or answer cues. We also tested 925 presentations from published sources: 802 from AcuityBench, 48 Levine vignettes, 65 Sen2Med symptom reports and 10 HELPMed scenarios (11–15). With their altered versions and decision questions, these formed 25,158 case-and-question entries. Across development and evaluation, 2,483,222 model calls were documented (**Table S9**).

Across constructed and published cases, automated testing followed defined rules for each task. We tested selected cases repeatedly after adding clinician advice, social labels, requests, refusals or repeated disagreement. All systems received the same inputs. We compared each altered version with its original and used neutral additions to measure changes attributable to wording alone. The main outcome was the difference between recommendation changes after a test addition and changes after a neutral addition. Where an applicable reference answer was available, we also assessed correctness. Repeatability was summarized descriptively. For Jev, it was the share of repeated identical requests that returned the first recorded answer, across 617,837 comparisons from 458,393 repeated inputs. For Decisions and Luna, we compared the three requests made for each entry: the three Decisions probability vectors were identical for all 25,145 entries with three complete responses, and Luna’s three answers agreed for 23,422 of 25,158 entries.

Constructed cases and earlier published benchmarks informed workflow development and threshold estimation. Models received the clinical text and questions without labels identifying the experimental condition. We then evaluated the selected workflow, with thresholds unchanged, on the 75 Sen2Med and HELPMed cases included in the published collection. Detailed testing rules, case selection, validation and reference standards are provided in **Supplementary Materials**.

### Paired workflow

Passage removal required an answer change, LLM classification of the passage as not reporting a patient finding, and a probability shift exceeding the 99th-percentile threshold estimated from unaltered messages. The classification used majority voting over repeated LLM readings, three in the fixed external test and in the Decisions team. Otherwise, the original answer was retained. The emergency check required an original emergency probability of at least 0.30 and emergency advice in all three LLM readings of the selected text. It could only raise urgency. We counted passage removals separately on unaltered and altered messages. Alternative workflows tested during development are described in **Table S18. Supplementary Materials** give the deletion rules, probability calculations and thresholds.

### Outcomes and analysis

Raw decision change was the percentage of altered inputs that changed the same system’s answer to the unaltered case. We calculated excess decision change by subtracting the neutral-control change rate from the test-addition change rate. For example, rates of 30% and 5% yield 25 additional changes per 100 comparisons.

Comparisons required usable responses from all five systems and matched neutral responses where applicable. The Decisions team returned usable outputs for 24,848 of 25,158 entries; 310 were unavailable because of refusals and were not imputed. Related presentations and versions were grouped by source scenario, and test sets were analyzed separately. **Supplementary Tables S8 and S9** report case selection, reuse and analysis denominators.

Overall summaries gave each challenge family equal weight within its test set, including stable controls. The descriptive group comparison in **Figure 1** also weighted the two decision models and their two teams equally. We used source-scenario bootstrap resampling for 95% confidence intervals: 10,000 resamples for individual outcomes and reference agreement, and 20,000 for family averages and paired reductions. Intervals were exploratory and unadjusted for multiple comparisons. **Supplementary Materials** describe the resampling and sensitivity analyses.

### Latency and cost

We summarized recorded latency and cost descriptively, without new model calls. For single calls, we used the saved requests for all 802 original AcuityBench presentations, one per presentation for Jev and three each for Decisions and Luna, and report the median request time and the mean cost per 1,000 calls (**Table S17**). Jev and Luna were called through OpenRouter on 5 October 2026, and Decisions directly through OpenAI on 11 October 2026. Costs used the posted input rates of US$0.042 per million tokens for Jev (2), and US$0.10 for Decisions (3), and the provider-reported amounts for Luna.

For the paired workflow, we counted every documented call of the Decisions team: 361,616 calls, 354,059 to the decision model and 7,557 to the LLM, with a median of 12 probes per entry (interquartile range 10 to 14). Costs were calculated from recorded tokens at list prices. LLM readings of original texts and classifier votes were reused from the standalone runs; adding all of Luna’s standalone answers and the shared classifier votes at their recorded costs gives an upper bound for deployment. A single probe took a median of 0.26 seconds and a single LLM call 1.1 to 1.8 seconds; the time of a whole paired decision was not measured, because each step ran separately across all cases. The full run of 25,158 entries cost US$14.15 (US$0.56 per 1,000 entries), and at most US$0.77 per 1,000 with the reused readings added. Luna’s own three answers per entry cost US$0.19 per 1,000 entries, from its mean single-call cost (**Table S17**). We priced the same three answers with GPT-6 Sol and GPT-6 Astra by scaling that cost by OpenAI’s posted price ratios, 20-fold and 100-fold, giving at least US$3.7 and US$18.6 per 1,000 entries (16); because these models may generate longer outputs than Luna, these are lower bounds.

## Data Availability

The data supporting the analyses are provided in the manuscript and supplementary files. Constructed scenarios, model decisions, comparison mappings, analysis code and prompts are publicly available at https://github.com/MahmudOmar11/decision-only-medical-ai. Published benchmark data are available from their original sources, listed in the repository, subject to their original licences.

https://anonymous.4open.science/r/acuity-bench-C9CD

https://github.com/ashwinra-code/gpt-health-eval

https://doi.org/10.1101/2023.01.30.23285067

https://github.com/abinithago/Sen2Med

https://github.com/am-bean/HELPMed

## Declarations

### Ethics and governance

This analysis used synthetic scenarios and previously published case materials. No participants were recruited for this study. The authors considered institutional review board approval unnecessary for this analysis.

### Author contributions

M.O. conceived and led the study, designed the workflow, assembled the analysis datasets, performed the analyses and drafted the manuscript. A.R., Y.B., A.G., J.B.K., M.A., O.R.B., A.S., B.S.G., R.E.A. and A.W.C. contributed to study design, data access, validation, interpretation or manuscript revision. E.K. and G.N.N. supervised the study, validated the methods and results, and revised the manuscript. A.T. and D.P. reviewed and edited the manuscript.

### Data availability

The published benchmark datasets are available from their original sources. Controlled scenarios, aggregate data and permitted case-level outputs are available to editors and reviewers during peer review and will be deposited in a public repository with a persistent identifier upon publication. Third-party case text and raw provider outputs will be shared where source licenses and terms permit.

### Code availability

Study code and prompts are available to editors and reviewers during peer review and will be released on GitHub, archived with a persistent identifier, upon publication. Jev and OpenAI Decisions are proprietary third-party services, and their model weights are not available to the authors.

