## Supplementary Information for "Decision-only medical AI: early evidence and pairing with language models"

### Supplementary Methods

#### *Study populations and decision questions*

We compared Jev and OpenAI Decisions, which return probabilities over specified decisions, GPT-6 Luna as the language model (LLM), and two teams pairing a decision model with the LLM. The final collection contained 2,134 distinct starting presentations: 1,209 constructed for this study and 925 from published sources. Published material comprised 802 AcuityBench presentations, 48 Levine vignettes, 65 Sen2Med symptom reports and 10 HELPMed scenarios. The constructed presentations were generated with Claude Opus 5.5, adapting the automated case-generation approach and examples from our previous studies (1,2,3). Two physicians from the study team jointly screened these cases for clinical plausibility and unintended label or answer cues. Table S9 distinguishes starting presentations, case-and-question entries and model calls.

Triage questions offered home monitoring, medical review within weeks, medical review within 24-48 hours or emergency care now. Other questions asked whether emergency care was needed, whether a mental health assessment was recommended, or whether a named investigation or treatment should be recommended now. The mental health question concerned assessment, not specialist referral. Original messages were compared with matched neutral additions and the specified clinical, social or advice-related alterations. Authority prompting denotes reported clinician advice; doctor advice and nurse-friend advice remain separate conditions.

Testing scripts applied defined question, alteration and control templates to eligible cases within each task. This allowed repeated comparisons across a batch of cases while varying the cue of interest. Task-specific rules differed across clinical questions and source datasets. For AcuityBench additions, eligibility required a first-person message with at least four sentence units and excluded the Semigran component. We sampled 300 eligible messages using a fixed random seed. Each contributed seven versions: original wording, a neutral sentence, doctor advice, nurse-friend advice, two assigned social labels and a neutral descriptor. The insertion position was held fixed within each case across the corresponding variants. Clinical-history additions and minimal changes to clinical findings were evaluated separately from social labels.

#### *Selection of Sen2Med and HELPMed cases*

The external evaluation used Sen2Med (4) and HELPMed (5). Of 195 Sen2Med AskDocs cases, 161 had the five required source versions and at least three visit ratings per version. An AI-assisted content screen selected 65 adult, first-person descriptions of current symptoms with sufficient clinical detail. Selection excluded overlap with earlier study cases and four incompatible assigned cues. Selected cases then independently reviewed by the research team (MO, BG and GNN) for applicability.

Each selected Sen2Med case contributed an original, a neutral sentence, doctor advice, nurse-friend advice, two assigned social labels and a neutral label, totaling 455 inputs. HELPMed contributed ten scenarios and ten third-person versions. Description matching required similarity of at least 0.95 and preserved numeric tokens. Source urgency of at least 3 identified four emergency and six nonemergency scenarios. Refusal-related missingness left 64 Sen2Med cases in matched five-system comparisons.

*Systems and sampling*

Jev used snapshot 1.13-20260917. The matched standalone comparator used the first original response, rather than an average of repeated Jev responses. The LLM used the majority of three categorical responses; option order resolved ties. Decisions averaged three complete probability vectors for each original input and used one response per unique probability probe. Both Decisions and the LLM service returned the model identifier gpt-6-luna. Their comparison therefore tests interfaces and workflows without establishing independent model families.

The LLM requests specified low reasoning effort and a 2,000-token output limit. Sampling used provider defaults without an explicit temperature or seed. Decisions requests also omitted explicit temperature and seed settings. The returned model identifier does not establish unchanged model weights across collection dates. Recorded individual-call latency and cost were summarized descriptively as detailed in Table S17.

*Influence-guided collaboration*

The decision model evaluated the original text and automated deletions at paragraph, sentence and smaller-span levels. Deletions preserved digit strings and age or sex phrases; smaller fragments were considered when deleting a complete sentence would violate those rules. A candidate had to change the decision. The LLM classified whether the removed text reported a patient finding, using repeated judgments and majority voting; the external evaluation used three readings. A candidate was eligible only when the majority classified the removed text as not reporting a patient finding. The prompts contained clinical text and decision questions without labels identifying the experimental condition.

The classifier asked: "Does this sentence report a finding about the patient: a symptom, a sign, a vital sign, an examination finding, a test result, or a medical condition, medication, allergy or procedure in the patient's history?" The extracted passage was presented as a sentence and required a yes or no answer. This classification identified patient findings; it did not establish whether the statement was true or sufficient to justify a decision.

Influence was the absolute change in cumulative probability across the urgency boundary crossed by a decision-changing deletion. Candidates were ranked by influence, with shorter spans preferred when influence was equal. For each decision question and original decision level, the threshold was the 99th percentile of the highest eligible influence among unaltered messages. Strata with fewer than 50 messages pooled within the question. A deletion was accepted only when its influence exceeded the threshold; otherwise the original decision was retained. The provisional team answer came from the decision-model probability vector for the selected text.

A separate emergency check could increase urgency. When the provisional answer was nonemergency, an original binary emergency probability of at least 0.30 and emergency advice in all three LLM responses on the selected text triggered an emergency recommendation. If no deletion was accepted, the LLM read the original text. This rule could escalate care but could not suppress emergency advice. Missing required classifier or selected-text responses remained missing.

The Jev and Decisions workflows used separate influence thresholds because their probability scales differed. Thresholds for controlled scenarios and each published benchmark were estimated during workflow development. The AcuityBench thresholds were transferred unchanged to Sen2Med and HELPMed; those external cases did not enter threshold estimation. Tables S10 and S11 report all decision-level thresholds.

*Clinical references*

AcuityBench and its addition cohort supplied source urgency labels or acceptable ranges (6). Component material included HealthBench (7) and symptom-checker cases (8). Ramaswamy vignettes used clinician-defined triage references that could permit adjacent urgency levels (9). Levine cases used the released source references (10). HELPMed supplied scenario emergency status, whereas Sen2Med distinguished care from home care and could not establish appropriate ER urgency. The Ramaswamy analysis used the expanded public release of 78 case formats from 39 clinical scenarios; the associated paper reported 60 formats from 30 scenarios.

Controlled-scenario references were specified during case construction or obtained through model-rater consensus, according to the task. Indicated-intervention cases were retained when two model raters agreed with the writer's answer. The physician screen assessed clinical plausibility and unintended cues; it did not provide independent adjudication of the target answer for every case or alteration. Reference sources and their application to altered cases are distinguished in Table S1.

We considered clinical relevance separately from agreement with a reference. Neutral additions and perspective changes retained the original reference when they preserved clinical findings. Advice, access barriers, refusal, preference, depression history and social context were not assumed to leave the appropriate action unchanged. Reassurance and demographic variants were inspected for changes to clinical facts. Variants without an applicable reference remained unclassified for directional accuracy analysis. To be noted clearly, the reassurance benchmark retained source references but lacked physician adjudication of altered facts.

*Decision changes, corrections and newly introduced errors*

For each system, decision change was the percentage of altered inputs that changed its own original answer. Where neutral controls were available, excess change was the difference between the cue-associated and neutral-control percentages. Thus, 25 percentage points represents 25 additional decision changes per 100 comparisons beyond neutral wording, with the stated within-case weighting. It is a difference between change rates, not an accuracy or harm score; values below zero indicate fewer changes after the cue than after neutral wording. An exact pair comprised an original input, its altered version, a decision question and a system. With an applicable reference, we classified each pair as agreement followed by disagreement, disagreement followed by agreement, agreement in both, or disagreement in both. Acceptable reference ranges were preserved.

Each team was also compared with its own standalone decision model on identical altered inputs. A corrected answer changed standalone disagreement with the reference to team agreement; a newly introduced error did the reverse. Prevention of an alteration-induced error required both original answers to agree with the reference and only the standalone answer to become discordant after alteration. We separately counted cases in which the team blocked a standalone correction. Loss of emergency advice required the same system to recommend emergency care for the original and to stop recommending it after alteration.

*Missing data and statistical analysis*

The retained comparison comprised 25,158 case-and-question records. The Decisions team returned usable outputs for 24,848; 310 were unavailable because of model refusals. Refusals were not resampled and missing decisions were not imputed. Exact-pair analyses required original and altered decisions for all five systems. Excess decision change also required neutral comparisons and could therefore have a different denominator from reference-agreement analyses. Table S8 reports the principal analysis populations.

We estimated uncertainty with 10,000 source-scenario percentile bootstrap samples. Formats and repeated versions from the same scenario were resampled together, retaining all systems within each sample. Each outcome retained its specified within-case weighting. Results were analyzed separately by source and reference definition. Intervals were exploratory and not adjusted for multiplicity. Comparisons with fewer than ten source scenarios were interpreted descriptively; intervals could not be estimated from a single scenario.

Sensitivity analyses restricted comparisons to complete neutral controls, single-level references or pairs for which both systems' original answers agreed with the reference. The companion data include the full outcome estimates, reference transitions, team comparisons and sensitivity analyses described in Table S12.

*Synthesis methods*

We used an exploratory family-level summary of each system's sensitivity. Within each family, excess change was the percentage of answers changed by test additions minus the percentage changed by neutral additions, using matched five-system responses. We averaged these family estimates with equal weights within each test set. The per-100 interpretation therefore describes an equally weighted mix of the tested challenge families, rather than a pooled case count. The weights do not represent the prevalence of these challenges in clinical practice. The relative reduction was the standalone-minus-team difference divided by the standalone average; it describes framing sensitivity, not the reduction in clinical errors.

For Figure 1's descriptive group summary, we also averaged the two tested decision-model configurations and their two teams with equal weights. Confidence intervals were calculated from these averages within the same joint source-scenario bootstrap resamples, preserving dependence between systems. The group summary describes the tested configurations; it does not pool clinical test sets or estimate a population of models.

The original controlled set included ten families: clinician advice in benign, ambiguous and emergency presentations; advice from others in ambiguous presentations; social labels in mental health assessment; refusal of an indicated intervention; demand for a nonindicated intervention; repeated disagreement in ambiguous and emergency presentations; and a clinical-history control. Stable and low-effect families were retained. The additional controlled set comprised relevance-screened labels, clinician advice and social labels. It reused some original cases and neutral responses and was analyzed separately. AcuityBench and Sen2Med each included doctor advice, nurse-friend advice and social labels. Comparisons without neutral controls were analyzed separately.

Each family retained its within-case weighting and complete-case rule. We resampled source scenarios jointly across systems and families, preserving each scenario's multiplicity wherever it reappeared. Families sharing scenarios were resampled together; disconnected source populations were sampled independently. AcuityBench formats were grouped by underlying scenario. The original controlled, additional controlled, AcuityBench and Sen2Med summaries included 428, 160, 280 and 64 source scenarios, respectively. These counts are not additive because some source material was reused.

We used 20,000 bootstrap resamples to obtain percentile 95% intervals for each average and standalone-minus-team difference. Positive differences indicate lower excess change with pairing. Intervals are pointwise and exploratory, without multiplicity adjustment. We examined raw and neutral changes separately, omitted each family in turn, and repeated the AcuityBench analysis with matched neutral-label controls (Tables S14-S16).

### Supplementary Results

Average excess change decreased for both teams in every test set. In the original controlled set, each of the seven families with a nonzero standalone effect had a lower team point estimate; the three stable controls remained stable. Relative reductions in the family averages were 79.7% for Jev and 82.3% for Decisions. Average reductions persisted after omitting any one family. In Sen2Med, the gain was driven mainly by doctor advice. Omitting that family left intervals including zero, and the Decisions social-label estimate increased (Tables S14-S16).

In the original controlled suite, mean excess change was 14.0 percentage points for the LLM, with a 95% CI of 12.9 to 15.3; 13.4 for the two decision-model configurations, with a 95% CI of 12.0 to 15.0; and 2.6 for their teams, with a 95% CI of 1.9 to 3.3. These descriptive averages cover the same ten families and 428 source scenarios as the individual-system estimates in Table S14.

Social-label effects varied across labels and systems. The Jev team's excess change for the unhoused label was -0.4 percentage points, compared with 93.5 for Jev alone. For the single-parent label, it remained 11.2 points, compared with 24.7 for Jev. Stable treated depression produced more change with the Jev team than with Jev alone, at 26.8 versus 20.3 points. These comparisons concern responsiveness to different information; assessment appropriateness was not adjudicated. Table S13 reports every social label and the clinical-history control.

In Sen2Med, doctor-advice excess change fell from 81.3 to 26.6 points for Jev and from 73.4 to 26.6 for Decisions. The nurse-friend effect remained high for Jev and its team, at 53.1 and 51.6 points, compared with 29.7 and 21.9 for Decisions and its team. These differences concern responsiveness to information whose clinical meaning can vary by case (Table S2).

Changes in reference agreement were mixed. On 294 neutral additions, the Jev team corrected six standalone reference disagreements and introduced seven; the Decisions team corrected one and introduced seven. In 279 intervention-refusal variants, the teams corrected 23 and 68 disagreements while introducing one and nine, respectively. These recommendation comparisons use the controlled scenarios' specified indication and do not adjudicate whether an intervention should be performed after refusal (Tables S3-S5).

All systems identified the four HELPMed emergencies. Decisions alone avoided emergency referrals in all six nonemergency scenarios, whereas its team added one. In the reassurance benchmark, Jev lost emergency advice on eight of 48 versions and its team on two; the six prevented losses arose from two underlying scenarios. Tables S6 and S7 report reference agreement and loss of emergency advice.

Reductions also appeared in raw decision changes. For Jev in AcuityBench, the average raw-change reduction was 19.9 percentage points and the excess-change reduction was 21.2 points; the difference reflected 1.4 points more neutral change with the team. Table S14 reports both components. Lower sensitivity alone does not establish that a removed influence was clinically inappropriate or that the resulting answer was correct.

### Supplementary Tables

#### Table S1. Clinical references and their application to altered cases.

| Task family | Reference | Altered-case use | Interpretation |
|---|---|---|---|
| AcuityBench neutral additions | Source urgency range | Invariant when no new finding | Source-referenced transition |
| HELPMed perspective | Scenario emergency status | Invariant wording change | Small descriptive check |
| Refusal of an indicated intervention | Writer and two model raters | Preference-sensitive | Agreement with specified indication |
| Ramaswamy reassurance variants | Clinician source range | Altered facts unresolved | Contextual benchmark only |
| Sen2Med original cases | Original-case care versus home label | Original cases only | Original-case agreement |
| Social labels and mental health assessment | None for appropriateness | No appropriateness reference | Association and stability only |

The reference appropriate to an original case may not remain appropriate after clinically meaningful information is added. Unclassified variants are retained in decision-sensitivity results but excluded from directional accuracy claims.

#### Table S2. Decision changes after advice, labels and clinical information.

Raw and neutral changes are percentages; excess change is their difference in percentage points. Intervals are exploratory 95% source-scenario bootstrap intervals. Each system is compared with its own original answer. Presentations from the same source scenario are clustered together. Study populations are analyzed separately. LLM denotes GPT-6 Luna.

*AcuityBench additions: doctor advice (293 units; 279 source cases)*

| System | Raw (%) | Neutral (%) | Excess (pp) | 95% CI |
|---|---|---|---|---|
| Jev | 70.0 | 2.7 | 67.2 | 61.6 to 72.9 |
| LLM | 62.5 | 5.1 | 57.3 | 51.4 to 63.3 |
| Decisions | 61.4 | 8.2 | 53.2 | 47.1 to 59.3 |
| Jev + LLM | 25.3 | 4.1 | 21.2 | 16.3 to 26.1 |
| Decisions + LLM | 25.6 | 7.5 | 18.1 | 13.1 to 23.2 |

*AcuityBench additions: advice from a nurse friend (293 units; 279 source cases)*

| System | Raw (%) | Neutral (%) | Excess (pp) | 95% CI |
|---|---|---|---|---|
| Jev | 46.8 | 2.7 | 44.0 | 38.2 to 49.8 |
| LLM | 20.5 | 5.1 | 15.4 | 10.5 to 20.2 |
| Decisions | 29.4 | 8.2 | 21.2 | 15.8 to 26.5 |
| Jev + LLM | 32.8 | 4.1 | 28.7 | 23.4 to 34.0 |
| Decisions + LLM | 22.5 | 7.5 | 15.0 | 10.1 to 19.9 |

*AcuityBench additions: social labels (292 units; 278 source cases)*

| System | Raw (%) | Neutral (%) | Excess (pp) | 95% CI |
|---|---|---|---|---|
| Jev | 9.6 | 2.7 | 6.8 | 4.1 to 9.9 |
| LLM | 6.3 | 5.1 | 1.2 | -1.2 to 3.6 |
| Decisions | 9.6 | 7.9 | 1.7 | -1.2 to 4.7 |
| Jev + LLM | 8.7 | 4.1 | 4.6 | 2.3 to 7.1 |
| Decisions + LLM | 9.8 | 7.2 | 2.6 | -0.7 to 5.8 |

*Sen2Med: doctor advice (64 units; 64 source cases)*

| System | Raw (%) | Neutral (%) | Excess (pp) | 95% CI |
|---|---|---|---|---|
| Jev | 85.9 | 4.7 | 81.3 | 71.9 to 90.6 |
| LLM | 81.3 | 3.1 | 78.1 | 67.2 to 87.5 |
| Decisions | 79.7 | 6.2 | 73.4 | 60.9 to 84.4 |
| Jev + LLM | 29.7 | 3.1 | 26.6 | 15.6 to 39.1 |
| Decisions + LLM | 34.4 | 7.8 | 26.6 | 14.1 to 39.1 |

*Sen2Med: advice from a nurse friend (64 units; 64 source cases)*

| System | Raw (%) | Neutral (%) | Excess (pp) | 95% CI |
|---|---|---|---|---|
| Jev | 57.8 | 4.7 | 53.1 | 40.6 to 65.6 |
| LLM | 34.4 | 3.1 | 31.2 | 20.3 to 43.8 |
| Decisions | 35.9 | 6.2 | 29.7 | 17.2 to 42.2 |
| Jev + LLM | 54.7 | 3.1 | 51.6 | 39.1 to 64.1 |
| Decisions + LLM | 29.7 | 7.8 | 21.9 | 10.9 to 34.4 |

*Sen2Med: social labels (64 units; 64 source cases)*

| System | Raw (%) | Neutral (%) | Excess (pp) | 95% CI |
|---|---|---|---|---|
| Jev | 3.9 | 4.7 | -0.8 | -6.2 to 4.7 |
| LLM | 9.4 | 4.7 | 4.7 | 0.0 to 10.9 |
| Decisions | 12.5 | 9.4 | 3.1 | -4.7 to 10.9 |
| Jev + LLM | 2.3 | 3.1 | -0.8 | -6.2 to 4.7 |
| Decisions + LLM | 14.8 | 7.8 | 7.0 | 0.0 to 14.8 |

*Controlled scenarios, assessment: social labels, grouped (250 units; 250 source cases)*

| System | Raw (%) | Neutral (%) | Excess (pp) | 95% CI |
|---|---|---|---|---|
| Jev | 47.0 | 1.3 | 45.7 | 42.5 to 48.7 |
| LLM | 82.9 | 6.1 | 76.8 | 72.3 to 80.9 |
| Decisions | 9.0 | 1.6 | 7.4 | 5.6 to 9.3 |
| Jev + LLM | 10.2 | 2.0 | 8.2 | 6.2 to 10.3 |
| Decisions + LLM | 9.4 | 2.8 | 6.6 | 4.5 to 8.7 |

*Controlled scenarios, intervention: patient refusal (53 units; 53 source cases)*

| System | Raw (%) | Neutral (%) | Excess (pp) | 95% CI |
|---|---|---|---|---|
| Jev | 15.5 | 1.0 | 14.5 | 8.0 to 21.6 |
| LLM | 11.3 | 0.0 | 11.3 | 6.8 to 16.2 |
| Decisions | 25.3 | 4.5 | 20.8 | 13.1 to 29.3 |
| Jev + LLM | 7.2 | 0.5 | 6.7 | 3.1 to 10.9 |
| Decisions + LLM | 3.0 | 0.4 | 2.6 | 0.8 to 4.9 |

**Table S3. Changes in agreement after neutral additions or perspective wording.**

*Neutral insertion (294 variants; 280 source cases)*

| System | New disagreement | Correction | Both agree | Both disagree |
|---|---|---|---|---|
| Jev | 4 | 1 | 219 | 70 |
| LLM | 5 | 4 | 213 | 72 |
| Decisions | 6 | 11 | 211 | 66 |
| Jev + LLM | 5 | 3 | 216 | 70 |
| Decisions + LLM | 7 | 7 | 209 | 71 |

*Neutral social labels (296 variants; 282 source cases)*

| System | New disagreement | Correction | Both agree | Both disagree |
|---|---|---|---|---|
| Jev | 6 | 2 | 217 | 71 |
| LLM | 6 | 4 | 212 | 74 |
| Decisions | 5 | 6 | 212 | 73 |
| Jev + LLM | 8 | 5 | 213 | 70 |
| Decisions + LLM | 7 | 7 | 209 | 73 |

*Perspective wording (10 variants; 10 source cases)*

| System | New disagreement | Correction | Both agree | Both disagree |
|---|---|---|---|---|
| Jev | 0 | 0 | 9 | 1 |
| LLM | 0 | 0 | 9 | 1 |
| Decisions | 0 | 0 | 10 | 0 |
| Jev + LLM | 0 | 0 | 9 | 1 |
| Decisions + LLM | 0 | 0 | 9 | 1 |

Values are counts of original-altered pairs classified against the applicable source reference. New disagreement denotes original agreement followed by disagreement; correction denotes the reverse. Cases recur across the two AcuityBench neutral conditions. HELPMed is a descriptive evaluation of ten scenarios.

**Table S4. Changes in recommendations after refusal of an indicated intervention.**

| System | New disagreement | Correction | Both agree | Both disagree |
|---|---|---|---|---|
| Jev | 41 | 0 | 204 | 34 |
| LLM | 30 | 0 | 249 | 0 |
| Decisions | 68 | 0 | 211 | 0 |
| Jev + LLM | 19 | 0 | 226 | 34 |
| Decisions + LLM | 6 | 2 | 268 | 3 |

All systems were evaluated on 279 variants from 57 source cases. Conditions were plain refusal, strong refusal, advice against intervention from a doctor, advice against intervention from an AI symptom checker and combined refusal statements. (Table S15).

**Table S5. Team corrections and newly introduced reference disagreements.**

*Neutral insertion (294 variants; 280 source cases)*

| Team | Corrected | New disagreement | Strict prevention / new error | Net change, pp (95% CI) |
|---|---|---|---|---|
| Jev + LLM | 6 | 7 | 1 / 2 | -0.3 (-2.8 to 2.0) |
| Decisions + LLM | 1 | 7 | 0 / 1 | -2.0 (-4.1 to -0.3) |

*Neutral social labels (296 variants; 282 source cases)*

| Team | Corrected | New disagreement | Strict prevention / new error | Net change, pp (95% CI) |
|---|---|---|---|---|
| Jev + LLM | 5 | 6 | 0 / 2 | -0.3 (-2.7 to 1.7) |
| Decisions + LLM | 3 | 5 | 0 / 2 | -0.7 (-2.7 to 1.0) |

*Indicated-test refusal (279 variants; 57 source cases)*

| Team | Corrected | New disagreement | Strict prevention / new error | Net change, pp (95% CI) |
|---|---|---|---|---|
| Jev + LLM | 23 | 1 | 23 / 1 | 7.9 (4.0 to 11.9) |
| Decisions + LLM | 68 | 9 | 68 / 6 | 21.1 (12.4 to 30.4) |

Each team is compared with its standalone decision model on the same altered input. Net change is the number corrected minus newly introduced disagreements, divided by all evaluated variants. Prevention and new-error counts in the strict comparison require both original answers to agree with the reference. Intervals use 10,000 paired source-scenario resamples and are exploratory.

**Table S6. Reference agreement on original cases.**

*AcuityBench primary (597 presentations; 559 source cases)*

| System | Reference agreement, n | Agreement (%) |
|---|---|---|
| Jev | 481 | 80.6 |
| LLM | 484 | 81.1 |
| Decisions | 492 | 82.4 |
| Jev + LLM | 487 | 81.6 |
| Decisions + LLM | 489 | 81.9 |

*AcuityBench ambiguous (190 presentations; 190 source cases)*

| System | Reference agreement, n | Agreement (%) |
|---|---|---|
| Jev | 112 | 58.9 |
| LLM | 115 | 60.5 |
| Decisions | 120 | 63.2 |
| Jev + LLM | 109 | 57.4 |
| Decisions + LLM | 118 | 62.1 |

*HELPMed (10 presentations; 10 source cases)*

| System | Reference agreement, n | Agreement (%) |
|---|---|---|
| Jev | 9 | 90.0 |
| LLM | 9 | 90.0 |
| Decisions | 10 | 100.0 |
| Jev + LLM | 9 | 90.0 |
| Decisions + LLM | 9 | 90.0 |

*Sen2Med any-care (64 presentations; 64 source cases)*

| System | Reference agreement, n | Agreement (%) |
|---|---|---|
| Jev | 51 | 79.7 |
| LLM | 53 | 82.8 |
| Decisions | 52 | 81.3 |
| Jev + LLM | 50 | 78.1 |
| Decisions + LLM | 52 | 81.3 |

Agreement is defined by each source's accepted answer or urgency range. AcuityBench primary and ambiguous-reference populations remain separate. HELPMed evaluates emergency versus nonemergency care; Sen2Med evaluates any care versus home care. These results concern original presentations.

Among 205 AcuityBench original presentations from 199 source scenarios with an emergency-only reference, Jev recommended emergency care on 173 and its team on 185. The separate emergency rule added 12 emergency recommendations. Passage removal added one and withdrew one, yielding no net change. Thus, the net increase of 12 arose from the emergency rule. This component analysis used the recorded workflow decisions.

**Table S7. Loss of emergency advice after reassuring information.**

| System | Emergency advice on original, n | Withdrawn after reassurance, n |
|---|---|---|
| Jev | 48 | 8 |
| LLM | 48 | 0 |
| Decisions | 46 | 1 |
| Jev + LLM | 48 | 2 |
| Decisions + LLM | 47 | 1 |

All systems were evaluated on 48 versions from 12 case formats and six source scenarios. A loss requires an emergency recommendation on that system's original response.

**Table S8. Analysis populations and missingness.**

| Population | Planned | Available | Analysis |
|---|---|---|---|
| All case-and-question records | 25,158 | 24,848 | Decisions-team output |
| AcuityBench primary originals | 607 | 597 | Five-system comparison |
| AcuityBench ambiguous originals | 195 | 190 | Five-system comparison |
| AcuityBench doctor advice | 300 | 293 | Matched neutral comparison |
| AcuityBench social labels | 300 | 292 | General-neutral comparison |
| Sen2Med cases | 65 | 64 | Five-system comparison |
| HELPMed original scenarios | 10 | 10 | Five-system comparison |
| Indicated-intervention refusal | 300 | 279 | Reference-agreement comparison |

The available population can differ by outcome because each comparison requires its original, altered and, where applicable, neutral responses. Counts refer to inputs or presentations, not independent patients. Source-scenario denominators appear with each result. The 312 combined symptom-framing and access-barrier comparisons lack an invariant altered-case reference and are excluded from directional reference comparisons.

**Table S9. Study inventory and external case selection.**

| Cohort | Records | Scope |
|---|---|---|
| Controlled clinical scenarios | 20,485 | Workflow development and clinical evaluation |
| AcuityBench | 802 | Workflow development and clinical evaluation |
| AcuityBench additions | 2,100 | Workflow development and clinical evaluation |
| Levine | 48 | Workflow development and clinical evaluation |
| Ramaswamy | 1,248 | Workflow development and clinical evaluation |
| External evaluation of fixed workflow | 475 | 455 Sen2Med and 20 HELPMed inputs; unchanged workflow |
| Sen2Med source AskDocs | 195 cases | 161 with complete source ratings; 65 selected after content and overlap screening |
| Sen2Med native rewrites | 260 versions | Excluded from study inputs |
| HELPMed source | 10 scenarios | Original plus third-person wording, 20 inputs |

Inventory rows describe case-and-question records unless source cases or scenarios are specified. The 20,485 constructed records, 4,198 published-benchmark records and 475 external-evaluation records total 25,158. The 75 external starting presentations are included in the 925 published-source presentations. Starting presentations were counted by distinct exact text, retaining alternative wordings and clinical versions while removing identical text duplicates. The 78 Ramaswamy formats are already included within AcuityBench and were counted once among starting presentations; reused source presentations were not recounted. Related presentations remained grouped by source scenario in the analyses. Source repository versions were 46bee75f for Sen2Med and 35b50de8 for HELPMed. Refusal-related missingness left 64 Sen2Med cases in matched analyses.

Across development and evaluation, the documented lower bound was 2,483,222 model calls. This comprised 2,119,753 historical response records, 1,853 logged additional attempts, 75,474 standalone Decisions calls and 286,142 new calls for the Decisions team, including recovery. These totals include repetitions, workflow probes and earlier model comparisons. Each call represented by both a start and a completion event was counted once. Some earlier transport attempts were not logged, so an exact total of all attempted or billed calls could not be reconstructed.

**Table S10. Triage influence thresholds.**

| Calibration cohort | Team | Level 0 | Level 1 | Level 2 | Level 3 |
|---|---|---|---|---|---|
| Controlled clinical scenarios | Jev + LLM | 0.2280 | 0.9229 | 0.6525 | 0.0840 |
| Controlled clinical scenarios | Decisions + LLM | 0.6800 | 0.6800 | 1.0000 | 0.3000 |
| Ramaswamy | Jev + LLM | 0.3645 | 0.3385 | 0.3956 | 0.2427 |
| Ramaswamy | Decisions + LLM | 0.4414 | 0.4414 | 0.3653 | 0.4886 |
| AcuityBench | Jev + LLM | 0.5844 | 0.5475 | 0.5818 | 0.8700 |
| AcuityBench | Decisions + LLM | 0.8200 | 0.9375 | 0.7089 | 0.8008 |
| AcuityBench additions | Jev + LLM | 0.5409 | 0.5238 | 0.4538 | 0.7120 |
| AcuityBench additions | Decisions + LLM | 0.7804 | 0.7804 | 0.6480 | 0.7327 |
| Levine | Jev + LLM | 0.4043 | 0.4043 | 0.4043 | 0.4043 |
| Levine | Decisions + LLM | 0.7978 | 0.7978 | 0.7978 | 0.7978 |
| Sen2Med and HELPMed | Jev + LLM | 0.5844 | 0.5475 | 0.5818 | 0.8700 |
| Sen2Med and HELPMed | Decisions + LLM | 0.8200 | 0.9375 | 0.7089 | 0.8008 |

Thresholds are absolute cumulative-probability changes across the crossed decision boundary. Triage levels 0, 1, 2 and 3 denote home care, review within weeks, review within 24-48 hours and emergency care. Influence must exceed the threshold strictly. Sen2Med and HELPMed use the AcuityBench thresholds without refitting.

**Table S11. Influence thresholds for other clinical decisions.**

| Decision | Original answer | Jev + LLM | Decisions + LLM |
|---|---|---|---|
| Hospital admission | 1 | 0.0000 | 0.0000 |
| Analgesia | 1 | 0.0000 | 0.0000 |
| Imaging | 1 | 0.0000 | 0.0000 |
| Specialist assessment | 1 | 0.0000 | 0.0000 |
| Emergency care indicated | 0 | 0.0318 | 0.3350 |
| Emergency care indicated | 1 | 0.0000 | 0.0802 |
| Mental health assessment | 0 | 0.0320 | 0.3559 |
| Mental health assessment | 1 | 0.0806 | 0.3553 |
| Intervention recommended now | 0 | 0.0630 | 0.0600 |
| Intervention recommended now | 1 | 0.3764 | 0.4381 |

Binary answers are coded 0 for no (not indicated) and 1 for yes (recommended). All tasks use the strict threshold rule: influence must exceed the cutoff. A zero cutoff therefore still requires positive influence. The separate original emergency-probability criterion is 0.30 for both teams.

**Table S12. Companion data guide.**

| Workbook sheet | Content |
|---|---|
| Data 1 | Decision change and reference-related outcomes |
| Data 2 | Reference eligibility and exclusions |
| Data 3 | Decisions-team paired differences |
| Data 4 | Original-to-altered decision transitions |
| Data 5 | Team corrections and newly introduced errors |
| Data 6 | Original-case agreement in matched comparisons |
| Data 7 | Original-case agreement in all available records |
| Data 8 | Sensitivity analyses |
| Data 9 | Transitions after refusal of indicated intervention |
| Data 10 | Team effects after refusal of indicated intervention |
| Data 11 | Loss of emergency advice |
| Data 12 | Outcome definitions |
| Data 13 | Influence thresholds |
| Data 14 | Study inventory |
| Data 15 | Average sensitivity by test set and system |
| Data 16 | Paired average reductions |
| Data 17 | Paired reductions by challenge family |
| Data 18 | Omission of each challenge family |
| Data 19 | Paired AcuityBench reference-agreement differences |

The companion workbook contains aggregate estimates and definitions. Blank outcome cells denote analyses without an applicable reference or estimable interval; zero denotes an observed value of zero. Point estimates are retained at full precision in the data. Abbreviations and comparison directions are defined on the Contents sheet.

**Table S13. Mental health assessment changes by social label and clinical control.**

| Label or control | LLM | Jev | Decisions | Jev + LLM | Decisions + LLM |
|---|---|---|---|---|---|
| Unhoused | 85.1<br>(80.5 to 89.5) | 93.5<br>(89.9 to 96.5) | 19.6<br>(14.5 to 24.9) | -0.4<br>(-1.6 to 0.8) | 9.6<br>(5.3 to 13.9) |
| Refugee | 85.1<br>(80.4 to 89.5) | 85.1<br>(80.4 to 89.6) | 5.6<br>(2.7 to 8.7) | 4.8<br>(1.9 to 8.0) | 9.2<br>(5.2 to 13.3) |
| Released from prison | 85.1<br>(80.4 to 89.3) | 75.1<br>(69.5 to 80.5) | 4.4<br>(1.6 to 7.5) | 6.8<br>(3.5 to 10.3) | 8.0<br>(4.4 to 11.7) |
| Undocumented immigrant | 73.9<br>(68.0 to 79.5) | 3.1<br>(0.7 to 5.7) | 2.0<br>(-0.1 to 4.3) | 4.4<br>(1.7 to 7.3) | 4.0<br>(1.1 to 6.9) |
| Sex worker | 83.1<br>(78.1 to 87.6) | 23.5<br>(18.1 to 29.1) | 10.0<br>(6.4 to 13.9) | 16.4<br>(11.9 to 21.2) | 3.6<br>(0.4 to 6.8) |
| Veteran | 79.1<br>(73.6 to 84.1) | 15.1<br>(10.5 to 19.9) | 1.6<br>(0.1 to 3.5) | 14.4<br>(10.1 to 18.9) | 3.2<br>(0.7 to 6.0) |
| Single parent | 46.3<br>(40.0 to 52.7) | 24.7<br>(19.3 to 30.3) | 8.4<br>(5.3 to 11.9) | 11.2<br>(7.2 to 15.6) | 8.4<br>(4.8 to 12.3) |
| Married parent | 8.3<br>(4.3 to 12.3) | 4.7<br>(1.9 to 7.9) | 0.0<br>(-0.9 to 1.1) | 4.0<br>(1.5 to 6.8) | 1.2<br>(-0.7 to 3.3) |
| Depression, stable on treatment | 83.9<br>(78.9 to 88.4) | 20.3<br>(15.2 to 25.5) | 10.4<br>(6.4 to 14.5) | 26.8<br>(21.1 to 32.7) | 9.6<br>(5.3 to 13.9) |

Values are excess assessment-recommendation changes beyond neutral wording, in percentage points. Each label was applied to the same 250 emergency notes; these are not separate patient cohorts. Stable treated depression is clinical history and married-parent status is a comparison condition. The task did not adjudicate assessment appropriateness. All neutral labels and individual-axis results are provided in Supplementary Data 1.

**Table S14. Average sensitivity with equal weighting of challenge families.**

Raw and neutral changes are percentages. Excess change is their difference in percentage points, with a 95% interval. Families receive equal weights within each set. Paired reductions compare each decision model with its team using the same 20,000 source-scenario resamples. Sets remain separate and share some source material.

*Original controlled suite*

| System | Raw change | Neutral change | Excess change (95% CI) |
|---|---|---|---|
| LLM | 16.7 | 2.7 | 14.0 (12.9 to 15.3) |
| Jev | 20.1 | 3.5 | 16.6 (14.8 to 18.3) |
| Decisions | 12.6 | 2.3 | 10.3 (8.6 to 12.1) |
| Jev + LLM | 5.9 | 2.5 | 3.4 (2.4 to 4.4) |
| Decisions + LLM | 3.4 | 1.6 | 1.8 (1.1 to 2.7) |

Paired reduction, pp (95% CI): Jev 13.2 (11.6 to 15.0); Decisions 8.5 (7.1 to 10.0).

*Additional controlled challenges*

| System | Raw change | Neutral change | Excess change (95% CI) |
|---|---|---|---|
| LLM | 61.8 | 2.9 | 58.8 (55.9 to 61.4) |
| Jev | 47.2 | 0.6 | 46.6 (44.9 to 48.4) |
| Decisions | 35.3 | 1.7 | 33.6 (32.1 to 35.0) |
| Jev + LLM | 4.2 | 0.8 | 3.4 (2.4 to 4.4) |
| Decisions + LLM | 3.5 | 2.2 | 1.3 (-0.3 to 2.8) |

Paired reduction, pp (95% CI): Jev 43.2 (41.0 to 45.4); Decisions 32.3 (31.0 to 33.6).

*AcuityBench*

| System | Raw change | Neutral change | Excess change (95% CI) |
|---|---|---|---|
| LLM | 29.8 | 5.1 | 24.6 (21.2 to 28.1) |
| Jev | 42.1 | 2.7 | 39.4 (35.4 to 43.3) |
| Decisions | 33.5 | 8.1 | 25.4 (21.4 to 29.3) |
| Jev + LLM | 22.3 | 4.1 | 18.2 (14.8 to 21.5) |
| Decisions + LLM | 19.3 | 7.4 | 11.9 (8.5 to 15.3) |

Paired reduction, pp (95% CI): Jev 21.2 (17.7 to 24.9); Decisions 13.5 (10.0 to 17.0).

*Sen2Med*

| System | Raw change | Neutral change | Excess change (95% CI) |
|---|---|---|---|
| LLM | 41.7 | 3.6 | 38.0 (31.8 to 44.3) |
| Jev | 49.2 | 4.7 | 44.5 (37.5 to 51.3) |
| Decisions | 42.7 | 7.3 | 35.4 (27.6 to 43.0) |
| Jev + LLM | 28.9 | 3.1 | 25.8 (19.0 to 32.6) |
| Decisions + LLM | 26.3 | 7.8 | 18.5 (10.9 to 26.0) |

Paired reduction, pp (95% CI): Jev 18.8 (14.6 to 23.4); Decisions 16.9 (10.4 to 23.7).

#### Table S15. Paired reductions by challenge family.

Values are standalone-minus-team differences in excess decision change, in percentage points, with exploratory 95% source-scenario bootstrap intervals. Positive values indicate lower sensitivity. Units retain within-family weights; cases are distinct source scenarios. A case can contribute several presentations. Zero-width intervals reflect no observed variation in these samples.

*Original controlled suite*

| Challenge | Units / cases | Jev reduction (95% CI) | Decisions reduction (95% CI) |
|---|---|---|---|
| Clinician advice, ambiguous cases | 160 / 80 | 31.6 (22.1 to 41.7) | 10.9 (4.6 to 18.0) |
| Clinician advice, emergency cases | 80 / 40 | 0.0 (0.0 to 0.0) | 0.0 (0.0 to 0.0) |
| Clinical history, emergency cases | 80 / 40 | 0.0 (0.0 to 0.0) | 0.0 (0.0 to 0.0) |
| Clinician advice, benign cases | 159 / 80 | 41.9 (38.9 to 44.7) | 45.9 (43.1 to 48.5) |
| Social labels, mental health assessment | 250 / 250 | 37.5 (33.8 to 41.1) | 0.8 (-1.7 to 3.3) |
| Advice from others, ambiguous cases | 160 / 80 | 8.5 (2.0 to 15.4) | 5.1 (1.0 to 10.0) |
| Refusal of an indicated intervention | 53 / 53 | 7.8 (4.0 to 12.1) | 18.2 (10.5 to 26.5) |
| Demand for a nonindicated intervention | 51 / 51 | 0.3 (0.0 to 1.0) | 1.3 (0.0 to 3.0) |
| Repeated disagreement, ambiguous cases | 158 / 80 | 4.4 (-0.6 to 10.3) | 2.5 (0.0 to 5.8) |
| Repeated disagreement, emergency cases | 80 / 40 | 0.0 (0.0 to 0.0) | 0.0 (0.0 to 0.0) |

*Additional controlled challenges*

| Challenge | Units / cases | Jev reduction (95% CI) | Decisions reduction (95% CI) |
|---|---|---|---|
| Labels screened for clinical relevance | 40 / 40 | 0.0 (0.0 to 0.0) | 0.0 (0.0 to 0.0) |
| Clinician advice, additional challenges | 160 / 40 | 97.5 (95.4 to 99.2) | 96.2 (93.0 to 98.9) |
| Social labels, additional challenges | 80 / 80 | 32.2 (25.8 to 38.5) | 0.8 (-1.6 to 3.5) |

*AcuityBench*

| Challenge | Units / cases | Jev reduction (95% CI) | Decisions reduction (95% CI) |
|---|---|---|---|
| Doctor advice | 293 / 279 | 46.1 (40.1 to 52.2) | 35.2 (28.9 to 41.4) |
| Nurse-friend advice | 293 / 279 | 15.4 (10.6 to 20.3) | 6.1 (1.4 to 10.8) |
| Social labels | 292 / 278 | 2.2 (0.0 to 4.7) | -0.9 (-3.5 to 1.7) |

*Sen2Med*

| Challenge | Units / cases | Jev reduction (95% CI) | Decisions reduction (95% CI) |
|---|---|---|---|
| Doctor advice | 64 / 64 | 54.7 (42.2 to 67.2) | 46.9 (34.4 to 59.4) |
| Nurse-friend advice | 64 / 64 | 1.6 (-3.1 to 6.2) | 7.8 (-1.6 to 18.8) |
| Social labels | 64 / 64 | 0.0 (0.0 to 0.0) | -3.9 (-11.7 to 0.8) |

#### Table S16. Sensitivity of set-specific pairing estimates.

Values are reductions in excess decision change, in percentage points, with 95% intervals. Omission rows recalculate the average after removing the named family. The clinical-history omission excludes the informative control. The AcuityBench row substitutes matched neutral-label controls for the general-neutral control. All family-omission estimates are included in the companion data.

| Analysis | Jev reduction (95% CI) | Decisions reduction (95% CI) |
|---|---|---|
| Original suite, omit benign clinician advice | 10.0 (8.2 to 12.0) | 4.3 (2.7 to 6.1) |
| Original suite, omit social labels | 10.5 (8.7 to 12.4) | 9.3 (7.8 to 11.0) |
| Original suite, omit clinical history | 14.7 (12.8 to 16.6) | 9.4 (7.9 to 11.1) |
| Additional suite, omit clinician advice | 16.1 (12.9 to 19.2) | 0.4 (-0.8 to 1.7) |
| Sen2Med, omit doctor advice | 0.8 (-1.6 to 3.1) | 2.0 (-4.7 to 8.2) |
| AcuityBench, matched neutral labels | 21.4 (18.0 to 25.0) | 14.1 (10.7 to 17.4) |

**Table S17. Recorded latency and cost of individual calls.**

| System | Saved calls | Latency, seconds (IQR) | Cost / 1,000 calls, US$ |
|---|---|---|---|
| Jev | 802 | 0.328 (0.308 to 0.369) | 0.0252 |
| OpenAI Decisions | 2,406 | 0.289 (0.212 to 0.315) | 0.0372 |
| GPT-6 Luna | 2,406 | 2.767 (2.266 to 3.325) | 0.0621 |

All 802 original AcuityBench presentations had complete successful records: one call each for Jev and three each for Decisions and Luna. Latency is the median of pooled individual calls, with interquartile range (IQR); cost is the mean multiplied by 1,000. These are individual-call resources, not resources for complete repeated-sampling or paired decisions. Jev costs were reproduced from recorded input tokens at US$0.042 per million; Decisions costs were calculated at US$0.10 per million. Luna costs used saved provider-reported amounts. Costs were not reconciled with invoices. Jev and Luna used OpenRouter on 5 October 2026; Decisions used OpenAI directly on 11 October 2026. Concurrency also differed. Times include the successful request round trip and response parsing but exclude local queueing, earlier failed transport attempts and retry delays. Additional probes, repeated judgments and processing for paired decisions are excluded. Comparisons are descriptive; no confidence intervals or significance tests were calculated.

**Table S18. Alternative workflows tested during development.**

| Workflow | Advice "go", benign (pp) | Social labels (pp) | Reference agreement , untouched benign (%) | Untouched decisions as Jev (%) | US$ per 1,000 | Seconds |
|---|---|---|---|---|---|---|
| Jev alone | 48.3 | 46.6 | 93.1 | 100.0 | 0.028 | 0.33 |
| Luna alone | 43.1 | 76.8 | 87.5 | 86.4 | 0.054 | 2.39 |
| Average of Jev and Luna | 43.4 | 74.4 | NR | 88.4 | 0.082 | 2.39 |
| Cascade: Luna decides when Jev is unsure | 45.1 | 68.2 | NR | 93.3 | 0.040 | 0.85 |
| Luna told to use clinical findings only | 4.4 | 18.6 | 88.1 | 87.5 | 0.064 | 2.34 |
| LLM cleaner, narrow, then Luna | 0.6 | 69.4 | NR | 85.9 | NR | NR |
| LLM cleaner, wide, then Luna | 0.2 | −1.1 | NR | 85.6 | NR | NR |
| LLM cleaner, narrow, then Jev | 0.0 | 43.3 | NR | 97.5 | NR | NR |
| LLM cleaner, wide, then Jev | 0.2 | −0.7 | 86.9 | 97.3 | 0.38 | 6.25 |

Development data from the team-design stage in early October 2026, before the final collection: 160 benign messages with added "go" advice from an own doctor, an advice-line nurse or a nurse friend, and 250 emergency department notes with social labels; Luna was the LLM. The first two value columns are excess decision changes beyond neutral sentences, in percentage points. Reference agreement is agreement with two blinded model raters on untouched benign messages, and untouched decisions as Jev is the share of control decisions identical to Jev alone. The narrow cleaner deleted opinions, advice, requests and tone; the wide cleaner also deleted social circumstances and group identity. The cleaner prompts and the clinical-findings instruction named the tested categories, so part of their gain is built in. Cost and time are per decision, one call per step. NR, not recorded for this arm. None of these workflows was selected; the final workflow is described in Methods.
